# Danger signs and management of suspected severe malaria cases at community level and in referral health facilities: an operational study in the Democratic Republic of the Congo

**DOI:** 10.1101/2021.11.30.21267082

**Authors:** Jean Okitawutshu, Aita Signorell, Jean-Claude Kalenga, Eric Mukomena, Giulia Delvento, Christian Burri, Fatou Mwaluke, Valentina Buj, Moulaye Sangare, Sylvie Luketa, Nina Brunner, Tristan Lee, Manuel Hetzel, Christian Lengeler, Antoinette Tshefu

## Abstract

**Background:** Evidence from one trial in Africa suggests that pre-referral Rectal Artesunate (RAS) can be a life-saving intervention for severe malaria in remote settings, where parenteral treatment is not available. Recognition of danger signs indicative of severe malaria is critical for prompt and appropriate case management.

**Methods:** An observational study was conducted in the Democratic Republic of the Congo (DRC) in the frame of the multi-country CARAMAL project, to assess the effectiveness of RAS under real-world conditions. Severely ill feverish children <5 years seeking care from a community-based healthcare provider were recruited in three rural health zones into a patient surveillance system. They were subsequently followed within the healthcare system and at home after 28 days to determine care seeking, antimalarial treatment provision and health outcomes.

**Results:** Overall, 66.4% of patients had iCCM general danger signs, as well as more specific danger signs. Children aged 2-5 years (aOR=1.58, 95% CI 1.20–2.08) and those presenting iCCM general danger signs were more likely to receive RAS (aOR = 2.77, 95% CI 2.04–3.77). Injectable treatment was less likely with RAS pre-referral treatment (aOR=0.21, 95% 0.13– 0.33). In the post-RAS phase, the case fatality ratio was 7.1%. Children not receiving RAS had a higher risk of dying, but this was not statistically significant (aOR = 1.50, 95% CI 0.86– 2.60). The odds of dying were reduced in patients without iCCM general danger signs, but just not statistically so (aOR = 0.64, 95% CI 0.38–1.06). Full oral therapy at a RHF was highly protective (aOR = 0.13, 95% CI 0.07-0.26), while a full treatment of severe malaria (injectable + oral) was shown to also decrease massively the odds of dying (aOR = 0.26, 95% CI 0.09– 0.79) compared to injectable treatment alone.

**Conclusions:** Better understanding the determinants of successful case management, and targeted improvements of the health system (especially the provision of a full course of an oral antimalarial) are crucial for improving health oucomes of children with suspected severe malaria.

## Background

In 2019, an estimated 229 million cases and 409,000 deaths due to malaria occurred worldwide, of which 93% and 94% in Africa (1). Severe malaria often leads to death or irreversible sequelae, if not appropriately treated. Prompt, effective antimalarial treatment coupled with quality supportive care can substantially reduce severe malaria mortality rates (2, 3). One of the major challenges remains the limited access to higher-level health facilities, where staff and equipment are available for the proper management of severe malaria cases. The situation is exacerbated for populations living in remote areas, resulting in treatment delays of several hours or even days (4, 5). Injectable artesunate is the first line treatment of severe malaria, both in children and in adults as compared to parenteral quinine (6-10). When delays in reaching referral health facilities (RHF) are expected, the World Health Organization (WHO) is recommending pre-referral treatment, either with parenteral antimalarial, or with a single rectal dose of 10mg per kilogram of body weight of artesunate (RAS) (2). RAS is also recommended as a pre-referral treatment in the integrated community case management (iCCM) guidelines (11, 12) or in primary health facilities (PHC) where injectable antimalarials are often not available (2, 13, 14). In many clinical settings, RAS was shown to be an excellent antimalarial, safe and well accepted (15, 16). It was also shown to be a saving-life intervention in a large three-countries randomized controlled trial (17). However, to-date is it unclear how much of that benefit can be obtained under real-world conditions (18).

The DRC has the second highest malaria mortality burden worldwide, with at least 45,000 deaths per year (1). Nearly all of its 81.3 million people (2016) are at a high risk of contracting malaria, with some variations of prevalence across the country, between rural and urban areas, and over small geographic distances (19-22). The landmass of the country is massive (over 2.1 million km^2^) and access to many areas is difficult. The national-level *Plasmodium falciparum* infection rate is still very high (45%), illustrating the hyper-endemic nature of malaria of the country (23). Likewise, the second Demographic and Health Survey (DHS-DRC II) found malaria prevalence rates ranging from 5% (North Kivu) to 38% in Orientale Province, with an average of 23% of children aged 6 to 59 months testing positive for malaria using microscopy (24). Similarly high values were obtained in another study (25), and even in the capital city of Kinshasa some urban peri-areas had community prevalence rates as high as 40% (19).

Although the country has improved both the prevention and case management of malaria in the recent decade (13, 26) and implemented the iCCM package widely, new interventions are urgently required to address the high number of preventable childhood deaths resulting from malaria. Much remains also to be achieved in better understanding the burden and patterns of severe fever illnesses at community level, as well as the patterns and circumstance of child deaths. Obviously, better dealing with severely ill children, who are at a high risk of dying, is of high priority for reducing the unacceptably high mortality in Congolese children. In some settings, the CFR for hospitalized severe malaria can be as high as 28% (27), which is well above the accepted CFR of below 10% in high quality care settings (7).

The results presented here are part of the Community Access to Rectal Artesunate for Malaria (CARAMAL) project carried out in DR Congo, Nigeria and Uganda to assess the public health value of RAS as a pre-referral treatment under real-world conditions [Lengeler *et al*., manuscript in preparation]. The main impact and operational study results for the three sites are presented elsewhere (28). The aim of the present work was to describe the distribution of signs and symptoms, including danger signs among children <5 years enrolled with an iCCM episode of severe febrile illness in the CARAMAL project. In a second step, we assessed the effect of key danger signs on main study outcomes: RAS use, Referrals, receiving injectable artesunate at a RHF, and key health outcomes including mortality.

## Methods

### Study site

This study was conducted in three rural Health Zones (HZ) in the western part of the DRC: Kenge located in Kwango Province, Ipamu and Kingandu located in Kwilu Province (Figure 1).

**Figure 1.**
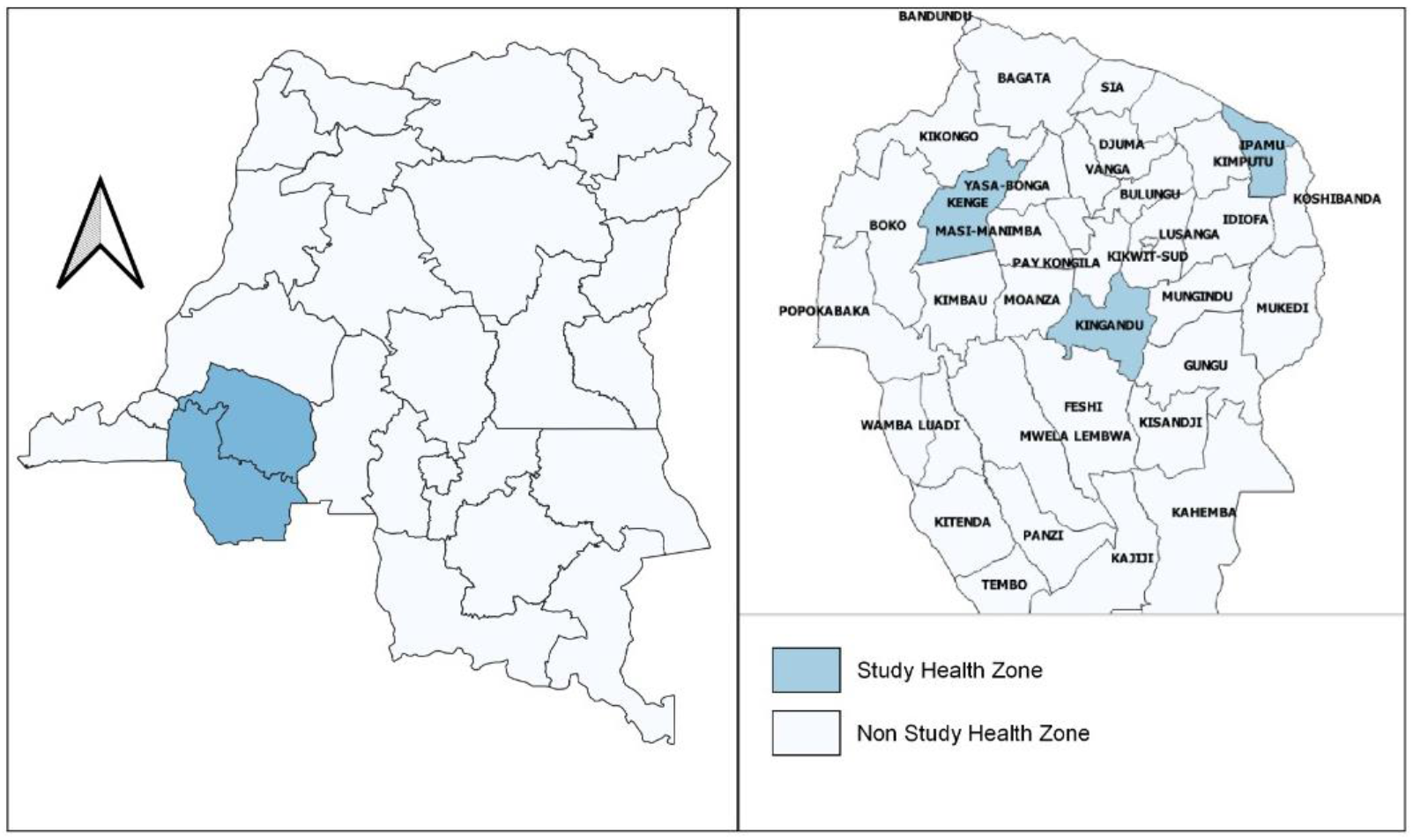
Map displaying the three CARAMAL study health zones in the Democratic Republic of the Congo.

The selection of the study areas was driven by scientific and operational considerations, including the population required for reaching the target sample size, a functioning iCCM programme supported by UNICEF, a functioning referral system, and an acceptable security situation. The three HZ have a combined population of approximately 785’968inhabitants, with an estimated 145’107children <5 years (https://www.worldpop.org, 2018). The peripheral level of care is composed of 42 functioning Community Health Care Sites (CHCS) and an extensive network of 152 Primary Health facilities (PHC) from the public, missionary and private sectors. The reference level of care comprises 19 Referral Health Facilities (RHF) including 16 Referral Health Centers and 3 General Referral Hospitals.

A CHCS is equipped with two community health workers (CHW) trained on iCCM algorithms, while nurses at PHCs adopt the Integrated Management of Childhood Illness IMCI strategy and provide a minimum package of preventive and curative care including the provision of RAS since severe cases can often not be managed at that level. By contrast, the vast majority of RHFs are staffed by at least one medical doctor and offer a much more comprehensive package of care, including blood transfusion as well as management of complications. Distances between facilities are often large, and there is no organized public transportation system in the study areas, so people mainly move by foot or bicycle.

### Study design

CARAMAL was an observational study based on a before-and-after plausibility design (29) in the framework of the RAS roll-out through established CHCS and PHCs. An extensive description of the study design and methods is available elsewhere [Lengeler *et al*., manuscript in preparation].

The core of the CARAMAL evaluation was a Patient Surveillance System (PSS) that was run for 10 months at baseline and 16 months after RAS introduction. The PSS allowed to enroll and track suspected severe febrile illness / severe malaria cases with regard to their clinical profile including danger signs and symptoms, their treatment pathway at community and RHF levels, and to determine treatment outcomes through a 28-days home visit.

Health care providers at all levels, including CHWs, PHCs and RHFs, underwent initial training sessions on the effective use of RAS according to the country’s iCCM guidelines, the diagnosis of malaria and severe case management. The training sessions were held prior to the RAS roll-out and were led by national, provincial and local health authorities, with the support of UNICEF. In addition, CARAMAL provided RAS to all CHWs and PHC facilities, and supplied injectable artesunate to referral facilities that were out-of-stock. The latter was the only other intervention in addition to RAS implemented by the project, so that the RAS evaluation could truly take place in real-world conditions, and hence the findings be generalizable.

### Participants

We enrolled all children below 5 years of age who were seeking care at a community-based CHW or PHC for a current severe febrile illness episode according to the following case definition: current fever or a recent history of fever, plus at least one iCCM danger sign indicating the need for referral to a higher-level health facility. In DRC, the danger signs identifying a child as eligible for referral and hence, RAS pre-referral treatment were: (1) vomiting everything, (2) convulsions, (3) not being able to drink/eat, and (4) being very sleepy or even unconscious. In addition, “unable to sit or stand up” and “general weakness / prostration” were also considered danger signs as per the local iCCM algorithm (collectively referred to as “*iCCM general danger signs*”). Of note is that these danger signs are not congruent with the WHO definition of severe malaria that is based on a more comprehensive clinical evaluation (2).

Children were eligible to enter the study and receive pre-referral RAS treatment if they fulfilled the inclusion criteria above, and if an informed consent from their caregiver was obtained (see specific section below). Children were ineligible if they were ≥5 years or had no permanent residence in the project areas. A second group of children was enrolled after directly attending a RHF, in order to describe comprehensively the epidemiology and management severe febrile illness / malaria. This group of children was obviously not eligible to receive RAS since they did not have to be referred, and hence are not considered here.

### Distribution of danger signs and symptoms

According to the WHO/UNICEF iCCM guidelines (12), there are four general danger signs: (1) convulsions, (2) unusually sleepy or unconscious, (3) not able to drink or feed anything and (4) vomits everything.

In the DRC iCCM guidelines, four groups of signs indicate a need for referral and should be systematically searched for by the CHW:

A. General danger signs, including (1) convulsions or history of recent convulsions, (2) unconscious or unresponsive to external stimuli, (3) not able to drink or breastfeed, and (4) vomiting everything. These are congruent with the WHO/UNICEF iCCM general danger signs.
B. Warning and severity signs, including “less than 2 months old”, “white palm”, “nutritional status RED”, “often ill”, “Illness lasting 14 days or more” or “fever lasting 7 days or more”, “very weak”, “difficulty breathing with severe chest indrawing or wheezing” and “becomes sicker despite adequate home care”.
C. Severe acute malnutrition signs: “visible and severe emaciation” and “edema of lower limbs”.
D. Others signs: “fever + generalized rash”, “blood in the stool” or “stool too liquid like water” and “diarrhea with dehydration”.

The formal recommendation to use pre-referral RAS and refer to a formal / higher level health facility is as follows: children aged 6 months to less than 6 years brought to a CHW or PHC with fever or history of recent fever (within the past 24 to 48 hours), plus at least one of the “general” danger signs listed under A above. However, in practice RAS was also administered (but not consistently) to children with warning and severity signs listed under (B-D) - except in “less than 2 months old”.

### Procedures

#### Records and referral

A child was provisionally enrolled on “day 0” at the peripheral level following its first contact with the health system. After a clinical assessment, the CHW or PHC health worker performed a malaria rapid diagnostic test (mRDT) for study purposes, and recorded the patient information in the individual patient file or in the patient logbook of the CHW or PHC. In a second step, the child was reported as a new case to the CARAMAL study nurse based at the nearest referral health facility (RHF). The child was then provisionally entered into the CARAMAL database, and a home visit was scheduled 28 days after the initial contact with the CHW or PHC. Reported data included demographic characteristics of the child such as its name, gender, weight, address/village, and name of the caregiver. The recorded information included as well the clinical status including current or history of fever, danger signs reported by the child’s caregiver, or found by the health worker during initial clinical assessment.

In addition, the date of the initial visit, as well as procedures and treatment provided including administration of RAS were reported. All children enrolled at peripheral level were subsequently referred to an identified RHF for severe disease management.

#### During admission (RHF)

A high percentage of children (67%) successfully completed referral to a dedicated RHF (30). At the RHF, the referred children were assessed by the health worker and treated according to the DRC national treatment guidelines (31). During admission at a RHF, trained CARAMAL study nurses extracted key patient information including case management information from facility records. This information was complemented by direct observation and communication with resident health workers and entered into an ODK Collect based electronic Case Report Form (https://opendatakit.org/). Data collected at this point included clinical assessment on arrival, test results, final diagnosis, treatment provided, daily clinical assessments, and condition of the child at discharge.

Local CARAMAL staff underwent intensive training on the study protocol, study procedures and ethics before the start of the study.

#### Day-28 follow-up home visits

##### Interviews

For all children enrolled by a community-based provider, home visits were conducted by the same CARAMAL study nurse between 28 and 30 days after initial enrollment. These home visits aimed to collect data on the child’s current health status and retrospectively record the history of fever, signs and symptoms, including RAS, the treatment seeking pathway during the past 28 days and antimalarial treatment received. This was done through an interview with the parent or caregiver, using a structured questionnaire. Furthermore, perception of RAS pre-referral treatment as well as parent’s/caregiver’s experience and attitudes towards the use of RAS were collected. Finally, some basic information on financial costs incurred by the caretakers during the disease episode were elicited. For deceased children an interview with the parent or guardian of the deceased child was postponed to 4 weeks after the date of death, to respect the grieving period. During the post-mortem interview, the CARAMAL study nurse elicited the circumstances and possible causes of death The ODK-based death form allowed collecting both written and audio recordings in the narrative section.

##### Blood testing

In addition to the interview conducted during the day-28 home visit, finger or heel-prick capillary blood was collected from all children for malaria antigen testing (CareStart HRP2 or HRP2/pLDH combined mRDT) and haemoglobin (Hb) level measurement (HemoCue Hb 201, Ängelholm, Sweden). Severe anemia was defined as Hb < 5 g/dl. Parents/caregivers of children who tested positive for malaria were advised to visit the nearest health facility for a full clinical evaluation and treatment in accordance with national guidelines. For children with danger signs or severe anaemia (Hb ≤5 g/dL), study staff brought them to the nearest referral health facility for treatment.

##### Data collection tools

we used structured electronic data collection forms designed on the Open Data Kit platform (ODK, https://opendatakit.org/) to capture data at each point of contact: at day 0, during admission in a RHF, and during the day-28 home visit. Each enrolled child was assigned a unique CARAMAL Identification number in order to link the data collected at different points in time and space. Completed data forms were monitored on a monthly basis by the local study supervisors in each health zone, and inconsistencies were immediately reviewed and subsequently corrected. Validated data were then uploaded on the online secure ODK Aggregate server hosted at the Swiss Tropical and Public Health Institute (Swiss TPH) in Basel, Switzerland. Study data was backed up daily.

### Study outcomes

The primary outcome of this study was the day 28 health status reported by the caregiver, defined as (i) healthy versus (ii) still sick versus (iii) deceased. Secondary outcomes included RAS administration defined as a binary variable (yes/no); referral completion to a dedicated RHF defined as binary variable (yes/no); injectable treatment at the RHF defined as a categorical variable (yes/no or not applicable for example if the child did not reach a RHF). Exposure variables of interest were the presence or not of iCCM general danger signs (yes/no). In addition, covariates of interest included enrolment location (CHW/PHC), study HZ, malaria test result on arrival at the RHF (positive / negative or not done), severe anaemia (hemoglobin (Hb) < 5g/dl), blood transfusion (yes/no), malaria oral treatment after parenteral treatment (yes/no), malaria test result on day 28 (positive / negative or not done), and anaemia (Hb < 11 g/dl) on day 28.

### Sample size calculation

The overall sample size of the CARAMAL multi-country study was estimated for CFR across the three project countries. We assumed the CFR to be 6% at baseline (historical CFR for severe malaria: 2.8% MATIAS Study DRC (8), 8.5% AQUAMAT (7)). A minimum of 6’032 severe malaria cases in children <5 years were required over 24 months to detect a 30% reduction in CFR between a 6 months baseline and 18 months following the roll-out of RAS, with 80% power and α = 0.05. In the case of our DRC specific analyses, we did not analyse specifically the impact of RAS on CFR. Given the large sample size required for the latter in each country, the sample size for the analysis presented here was largely sufficient.

### Statistical analysis

Data were downloaded as CSV files from the ODK aggregate online secure server using ODK-Briefcase version 1.13.1. The data were exported and analyzed in STATA version 16.0 (STATA Corporation, College Station, TX, USA). An Intention-to-Treat (ITT) analysis was done, which included all participants who were formally enrolled following informed consent, and for whom day-28 follow-up data were available. We computed the distribution of danger signs and symptoms among participants, stratified in pre-RAS and post-RAS periods, as well as RAS users and RAS non-users. Continuous variables were sumarized by their mean and standard deviation (SD), or median and interquartile range (IQR) when the distribution was skewed. Dichotomous outcomes were summarized as proportions, with 95% confidence intervals.

We used the Pearson Chi square test to compare percentages. In the first step, we assessed individual bivariate associations between outcomes and initial explanatory variables, using enrolling provider as random effect in univariable-mixed effects logistic regression. This provided crude odds ratios (Crude OR) with their 95% confidence intervals (95%CI), which guided the selection of variables to be included in subsequent multivariable models. In a second step, we built multilevel-mixed effects logistic regression models for each primary and secondary outcome and so adjusted for potential confounders. Again, we included enrolling provider as random effect to adjust for clustering at that level. Results are presented as adjusted odd ratios (aOR) and their 95%CI.

### Ethics

The CARAMAL study protocol was approved by the Research Ethics Review Committee of the World Health Organization (WHO ERC, No. ERC.0003008), the Ethics Committee of the University of Kinshasa School of Public Health (No. 012/2018), the Health Research Ethics Committee of the Adamawa State Ministry of Health (S/MoH/1131/I), the National Health Research Ethics Committee of Nigeria (NHREC/01/01/2007-05/05/2018), the Higher Degrees, Research and Ethics Committee of the Makerere University School of Public Health (No. 548), the Uganda National Council for Science and Technology (UNCST, No. SS 4534), and the Scientific and Ethical Review Committee of CHAI (No. 112, 21 Nov 2017). The study is registered on ClinicalTrials.gov (NCT03568344).

Consent was obtained in a two-step process given that the enrolled children were medical emergencies: a first provisional oral consent was obtained at the point of recruitment. The final written informed consent was then obtained during the first contact of the patient/caretaker with the study team - at the referral facility or during the day-28 home visit.

## Results

The study flow-chart (Figure 2) displays recruited study participants and their subsequent case management until their Day-28 outcome assessment. A total of 138 children were recruited directly at the RHF and were not analysed here.

**Figure 2.**
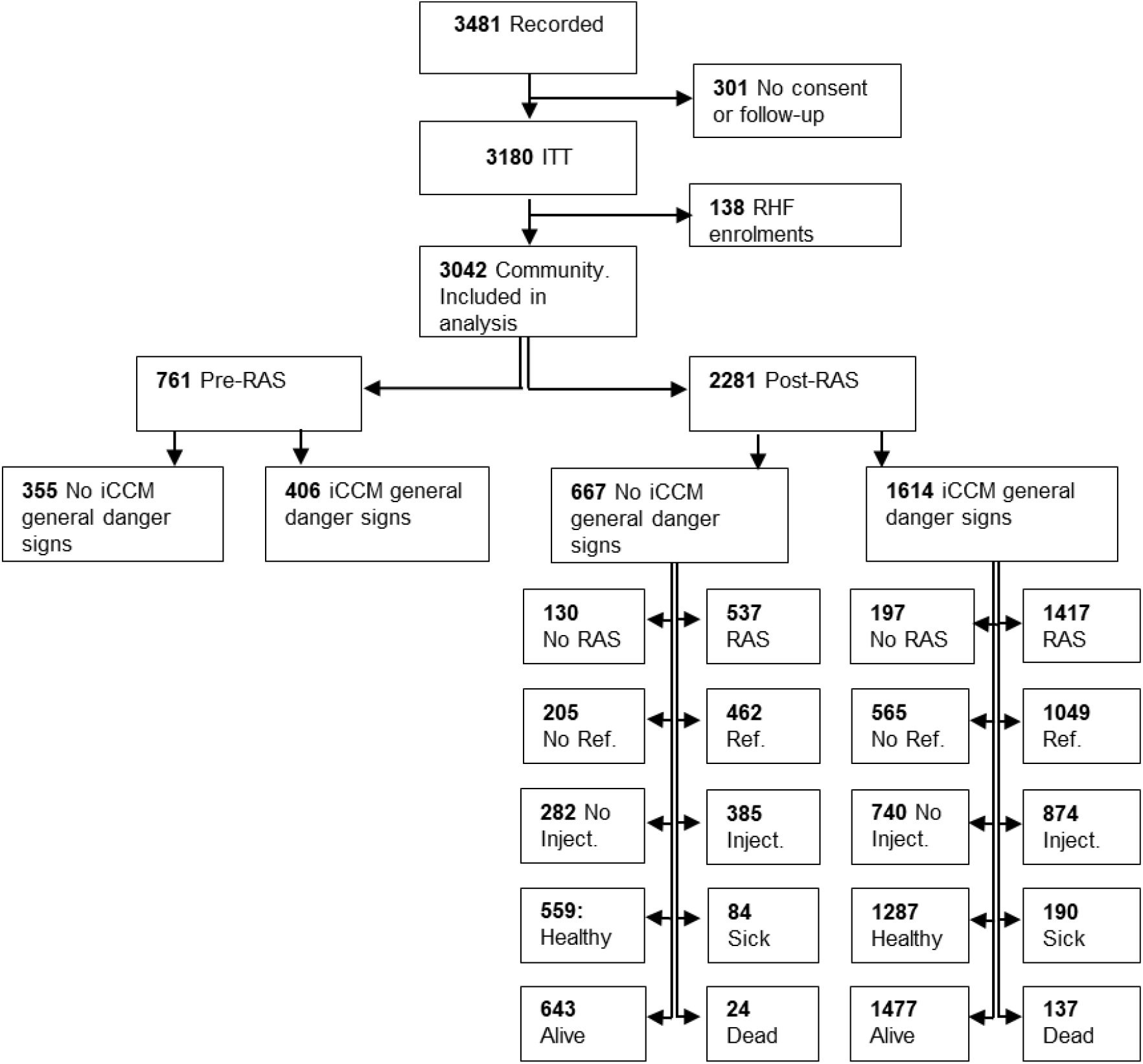
Study flow-chart. ITT = Intension-to-Treat. RHF = Referral Health Facility. iCCM = integrated Community Case Management. RAS = Rectal Artesunate. Ref. = Referral completed. Inject. = Injectable antimalarial treatment.

### Characteristics of study participants

Key characteristics of study participants are shown in Table 1. Between June 2018 and July 2020, a total of 3’042 feverish children <5 years old (median of age 2 years [IQR 1 - 3]) seeking care from a CHW or PHC provider were recruited into the study. Of those, 57.6% were children aged 0-2 years and 46.9% were female, with no difference in sex-ratio between the pre-RAS and post-RAS periods (p=0.93). Overall, in Kingandu HZ, significantly less children were recruited (813) compared to Kenge (1’101) and Ipamu (1’128) HZs. The vast majority of participants were enrolled at PHCs (94.6%) rather than by CHWs.

**Table 1.**
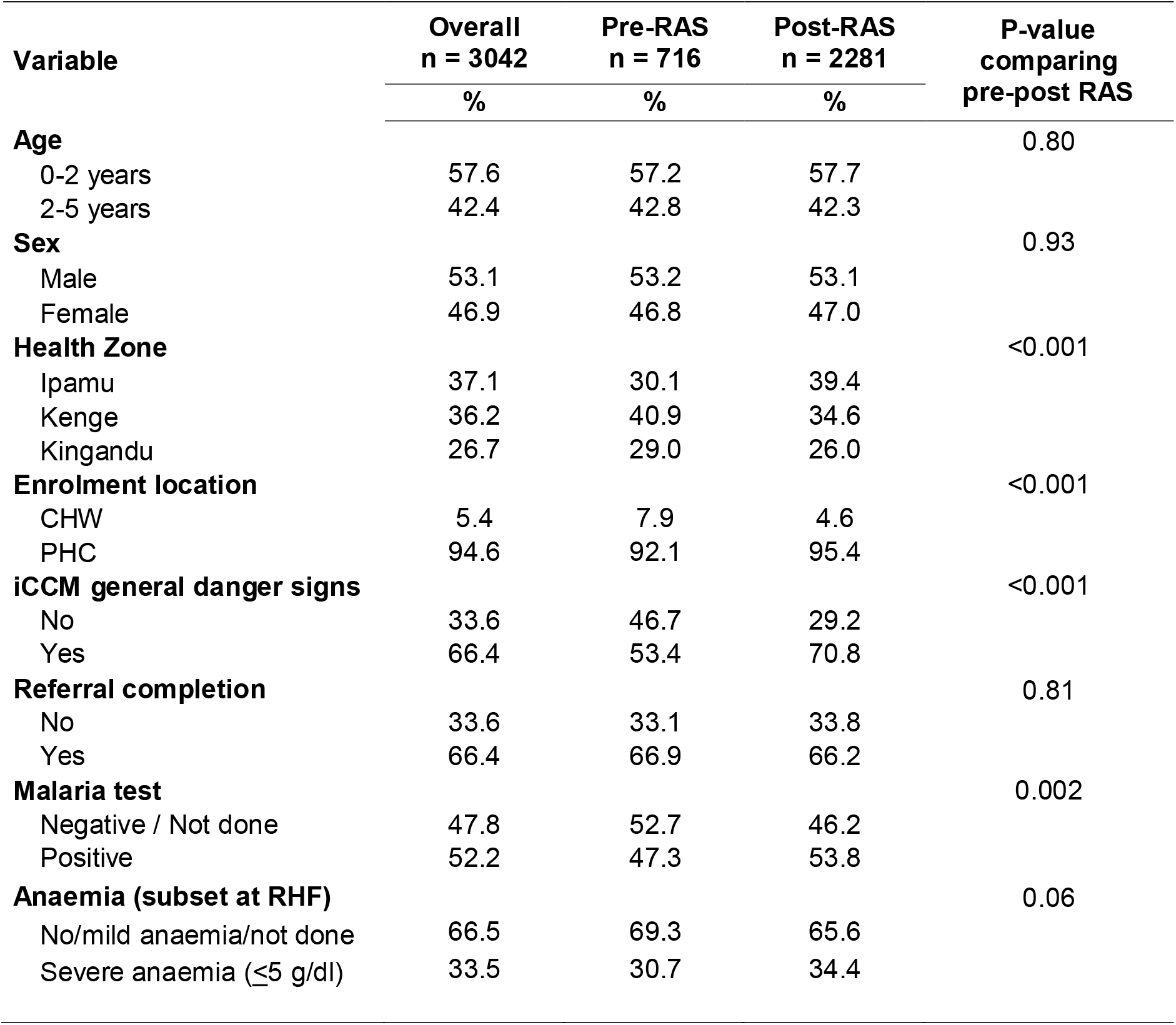
Characteristics of study participants at enrolment, by study phase. CHW = Community Health Worker. PHC = Primary Health Care. RHF = Referral Health Facility. RAS = Rectal Artesunate. iCCM = integrated Community Case Management.

Nearly two-thirds of patients (66.4%) presented iCCM danger signs upon enrollment (these signs are listed in Table 2). This proportion was only 53.4% during the pre-RAS period and rose markedly to 70.8% during the post-RAS period (p<0.001). Slightly more than half of the patients (52.2%) were tested positive for malaria at recruitment, with a higher proportion post-RAS (47.3% vs. 53.8%, *p*=0.002). This low proportion is due to the fact that iCCM and IMCI algorithms were used initially, and not the severe malaria case definitions. Only 66.4% of patients successfully completed referral to a dedicated RHF, without change between the pre-RAS and post-RAS periods (p=0.814). About 1/3 of the patients (33.5%) were anemic on arrival at RHF, with no significant difference observed between study phases (p=0.064).

**Table 2.** Danger signs and symptoms among children <5 years recruited at community level, by study phase. iCCM = integrated Community Case Management. RAS = Rectal Artesunate.

| Variable | Overall<br>n = 3042<br>% | Pre-RAS<br>n = 716<br>% | Post-RAS<br>n = 2281<br>% | P-value<br>comparing<br>pre-post RAS |
| --- | --- | --- | --- | --- |
| **iCCM general danger signs** |  |  |  |  |
| Convulsions | 40.8 | 30.4 | 44.2 | <0.001 |
| Not able to breastfeed, drink or eat anything | 36.2 | 31.3 | 37.8 | 0.001 |
| Unusually sleepy or unconscious | 18.9 | 23.7 | 17.3 | <0.001 |
| Vomiting everything | 8.5 | 8.9 | 8.3 | 0.58 |
| **Additional local iCCM danger signs** |  |  |  |  |
| Unable to sit or stand up | 26.1 | 9.7 | 31.6 | <0.001 |
| Difficulty in breathing | 9.2 | 11.2 | 8.5 | 0.03 |
| White palms or soles | 6.8 | 6.2 | 7.0 | 0.43 |
| Blood in stool | 1.5 | 2.2 | 1.2 | 0.05 |
| **Other Signs and symptoms** |  |  |  |  |
|  | **N = 2834** | **N = 703** | **N = 2131** |  |
| Weakness | 17.4 | 16.4 | 17.7 | 0.43 |
| Fever | 14.7 | 13.5 | 15.1 | 0.30 |
| Shivering | 7.7 | 13.5 | 5.8 | <0.001 |
| Nausea | 2.2 | 3.3 | 1.9 | 0.03 |
| Headache | 1.8 | 3.6 | 1.3 | <0.001 |
| Stomach ache | 1.7 | 2.2 | 1.5 | 0.20 |
| Cough | 1.6 | 1.7 | 1.5 | 0.70 |

The distribution of “standard” iCCM danger signs, as listed in (12, 32), as well as other danger signs used in the DRC context among study participants is shown in Table 2. “Convulsion” was the most frequent danger sign reported (40.8%). Compared to pre-RAS, a significantly higher proportion of children presented with convulsions or a history of convulsion during the post-RAS phase (p<0.001). “Not able to breastfeed, drink or eat anything” was frequently reported among study participants (36.2%) with a higher proportion recorded during the post-RAS phase (p=0.001). Although “unusually sleepy or unconscious” was reported in 18.9% of the children recruited, the baseline proportion (23.7%) was significantly higher than post-RAS (17.3%), p<0.001. “Vomiting everything” was the least frequent among iCCM danger signs, reported in less than 10% of recruited children, and with no significant difference between pre-RAS and post-RAS study phases. Among non-general iCCM danger signs, “unable to sit or stand up” was most frequently reported (26.1%), with a higher proportion during post-RAS phase (p<0.001). “Difficulty in breathing” was also reported more frequently during the post-RAS phase (p=0.03). The other signs “white palms or sole” and “blood in stool” were much less frequent.

Finally, Table 2 lists also a number of other signs and symptoms. Other danger signs not leading to RAS administration were also reported, but are not shown here because they were very infrequent and not relevant for the CARAMAL study: “fever for 7 days or more”, “cough for 14 days or more”, “diarrhea for 14 days or more”, “yellow eyes or jaundice”, “coke colored urine” and other uncommon symptoms including cough with sputum, watery diarrhea, skin rash and body or joint pains.

The results that follow include the use of RAS, and are therefore restricted to patients enrolled during the post-RAS phase of the study.

#### Determinants of RAS use

The contribution of different predictors associated with RAS use at CHW and PHC level is shown in Table 3. Based on these results, sick children aged 2-5 years were more likely to receive RAS compared to those aged 0-2 years (aOR = 1.58, 95% CI 1.20–2.08). There was no evidence of significant association between RAS use and gender or enrolment location. Significant heterogeneity in RAS use was observed among the three health zones. Compared to Ipamu as reference HZ and Kenge HZ (aOR = 0.69, 95% CI 0.41–1.18), RAS was significantly less likely to be used in the Kingandu HZ (aOR = 0.48, 95% CI 0.28–0.84). RAS use was conditional on the presence of at least one iCCM danger sign; consequently, children with iCCM general danger signs (category A) were significantly more likely to receive RAS (aOR = 2.77, 95% CI 2.04–3.77). Regarding the two additional signs triggering RAS use in DRC, the odds of receiving pre-referral RAS increased in both groups, while the association was significant in those “unable to sit” (aOR = 2.06, 95% CI 1.12–3.80), no such association observed in children suffering from asthenia (weakness) (aOR = 1.19, 95% CI 0.64–2.19).

**Table 3.** Determinants of RAS use by peripheral health workers. N = 2281. OR = Odds Ratio. CHW = Community Health Worker. PHC = Primary Health Care. 95% CI = 95% confidence interval.

| Determinant | n | % | Adjusted OR | 95% CI | p-value |
| --- | --- | --- | --- | --- | --- |
| **Age** |  |  |  |  |  |
| 0–2 years | 1316 | 57.7 | Ref. |  |  |
| 2–5 years | 965 | 42.3 | 1.58 | 1.20–2.08 | 0.001 |
| **Sex** |  |  |  |  |  |
| Male | 1210 | 53.0 | Ref. |  |  |
| Female | 1071 | 47.0 | 1.02 | 0.79–1.31 | 0.90 |
| **Enrolment location** |  |  |  |  |  |
| CHW | 104 | 4.6 | Ref. |  |  |
| PHC | 2177 | 95.4 | 0.87 | 0.40–1.89 | 0.72 |
| **Health Zone** |  |  |  |  |  |
| Ipamu | 899 | 39.4 | Ref. |  |  |
| Kenge | 790 | 34.6 | 0.69 | 0.41–1.18 | 0.17 |
| Kingandu | 592 | 26.0 | 0.48 | 0.28–0.84 | 0.01 |
| **Danger signs** |  |  |  |  |  |
| No/Others | 415 | 18.2 | Ref. |  |  |
| Yes (iCCM general) | 1614 | 70.8 | 2.77 | 2.04–3.77 | <0.001 |
| Weakness (asthenia) | 103 | 4.5 | 1.19 | 0.64–2.19 | 0.58 |
| Unable to sit | 149 | 6.5 | 2.06 | 1.12–3.80 | 0.02 |

#### Determinants of referral completion

Predictors associated with referral completion are presented in Table 4. Children in the age group of 2 to 5 years were significantly less likely to complete referral to a RHF (aOR = 0.71, 95% CI 0.54–0.93) than younger children. Compared to children enrolled by a CHW, PHC enrolments were associated with higher odds of completing referral (aOR = 4.22, 95% CI 1.09–16.32). Referral completion rates appeared lower in Kenge and Kingandu compared to Ipamu HZ, but a statistically significant decrease was only observed for Kenge HZ (aOR = 0.10, 95% CI 0.03–0.29).

**Table 4.**
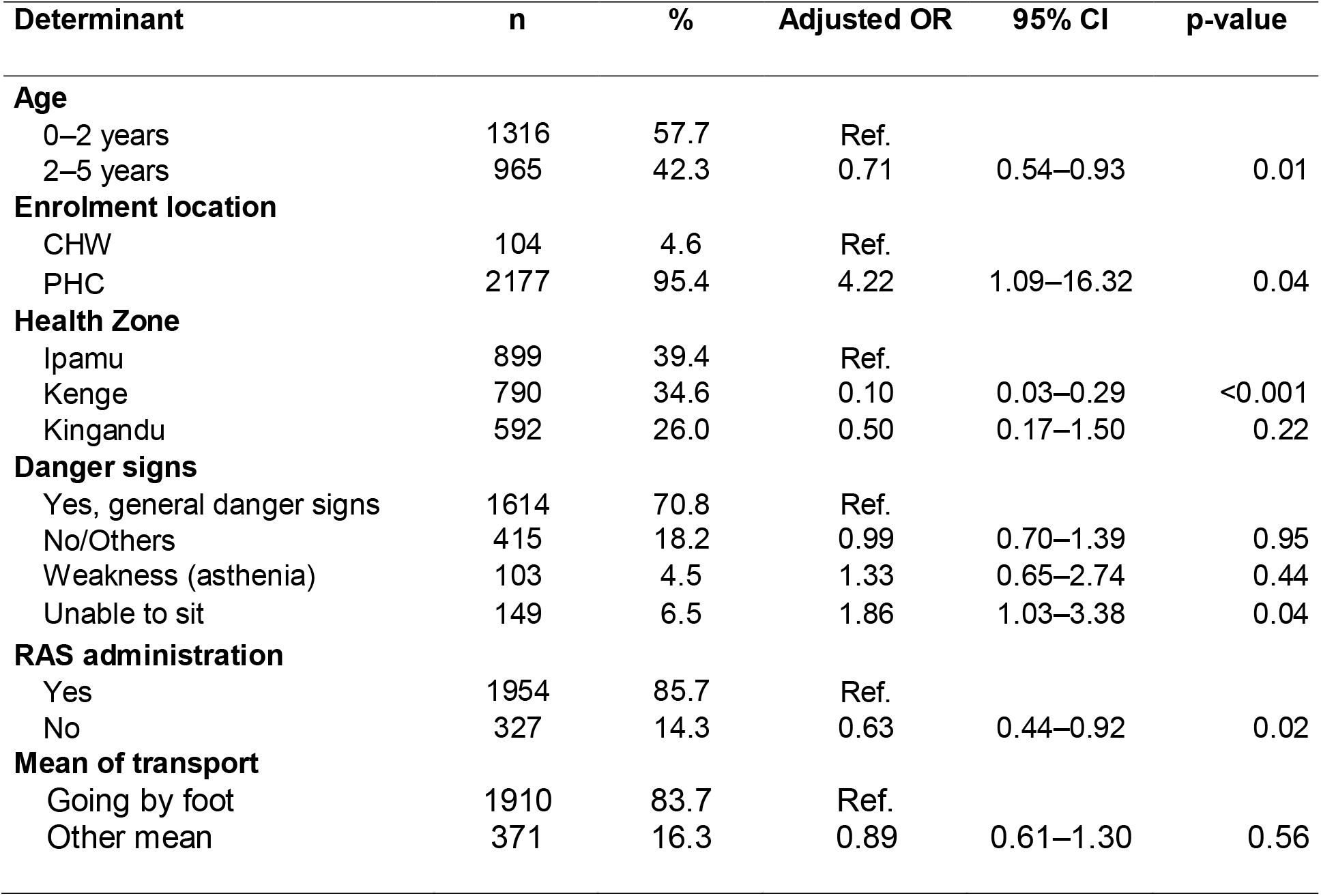
Estimated associations between selected determinants and referral completion. N = 2281. OR = Odds Ratio. CHW = Community Health Worker. PHC = Primary Health Care. 95% CI = 95% Confidence Interval. RAS = Rectal Artesunate. Ref.= Reference.

| Determinant | n | % | Adjusted OR | 95% CI | p-value |
| --- | --- | --- | --- | --- | --- |
| **Age** |  |  |  |  |  |
| 0–2 years | 1316 | 57.7 | Ref. |  |  |
| 2–5 years | 965 | 42.3 | 0.71 | 0.54–0.93 | 0.01 |
| **Enrolment location** |  |  |  |  |  |
| CHW | 104 | 4.6 | Ref. |  |  |
| PHC | 2177 | 95.4 | 4.22 | 1.09–16.32 | 0.04 |
| **Health Zone** |  |  |  |  |  |
| Ipamu | 899 | 39.4 | Ref. |  |  |
| Kenge | 790 | 34.6 | 0.10 | 0.03–0.29 | <0.001 |
| Kingandu | 592 | 26.0 | 0.50 | 0.17–1.50 | 0.22 |
| **Danger signs** |  |  |  |  |  |
| Yes, general danger signs | 1614 | 70.8 | Ref. |  |  |
| No/Others | 415 | 18.2 | 0.99 | 0.70–1.39 | 0.95 |
| Weakness (asthenia) | 103 | 4.5 | 1.33 | 0.65–2.74 | 0.44 |
| Unable to sit | 149 | 6.5 | 1.86 | 1.03–3.38 | 0.04 |
| **RAS administration** |  |  |  |  |  |
| Yes | 1954 | 85.7 | Ref. |  |  |
| No | 327 | 14.3 | 0.63 | 0.44–0.92 | 0.02 |
| **Mean of transport** |  |  |  |  |  |
| Going by foot | 1910 | 83.7 | Ref. |  |  |
| Other mean | 371 | 16.3 | 0.89 | 0.61–1.30 | 0.56 |

Compared to patients who had an iCCM general danger sign, having no danger sign did not seem to impact referral completion. No evidence of association was observed between referral completion and “weakness”, but children who were “Unable to sit” were more likely to complete referral (aOR = 1.86, 95% CI 1.03–3.38). Importantly, patients who did not receive RAS were significantly less likely to complete referral (aOR = 0.63, 95% CI 0.44–0.92). Finally, using other mean of transport including bicycle, motorbike, car … did not show a significant association with referral completion (aOR = 0.89, 95% CI 0.61–1.30) compared to those that went by foot as mean of transport.

#### Determinants of injectable treatment provision at RHF

The results are centered on 1511 children that completed referral successfully and thus would be provided injectable treatment while admission. Table 5 displays predictors associated with the provision of injectable antimalarial treatment for severe malaria in RHFs. There was no evidence of association between the provision of injectable antimalarial treatment and age of children or enrolment location. Considering Ipamu as reference HZ, injectable treatment was significantly more likely to be used at Kenge (aOR = 6.30, 95% CI 3.30–12.05) while no association was observed at Kingandu (aOR = 0.83 95% CI, 0.48–1.44). Patients who had iCCM general danger sign, those who had “asthenia” and those who were “unable to sit” did not show any evidence of association with the provision of injectable treatment at the RHF compared to those that had no iCCM general danger signs. Patients not treated with RAS at community level were less likely to receive injectable treatment at RHF (aOR = 0.21, 95% CI 0.13–0.33) compared to those that received RAS. Completing referral the same day or one day later was not significantly associated with increased odds of injectable antimalarial treatment provision compared to those that completed referral later (or with undocumented referral status (aOR = 0.95, 95% CI 10.65–1.41). A negative malaria test was logically very much associated with decreased odds of injectable treatment provision (aOR = 0.07, 95% CI 0.04–0.11). A significant decrease of antimalarial injectable provision was also found with those who did not have severe anaemia (aOR = 0.44, 95%CI 0.27–0.72). The presence of other comorbidities than anaemia was significantly associated with the use of injectable treatment (aOR = 2.36, 95% CI 1.62–3.44). Patients that did not receive a blood transfusion were less likely to be treated with injectable antimalarials (aOR = 0.53, 95% CI 0.32–0.87).

**Table 5.** Determinants of injectable antimalarial treatment for severe malaria at referral health facilities in community enrolments. N = 1511. OR = Odds Ratio. CHW = Community Health Worker. PHC = Primary Health Care Centers. RHF = Referral Health Facilities. RAS = Rectal Artesunate. 95% CI = 95% Confidence Intervals.

|  | n | % | Adjusted OR | 95% CI | p value |
| --- | --- | --- | --- | --- | --- |
| **Age** |  |  |  |  |  |
| Children (0–2 years) | 921 | 61.0 | Ref. |  |  |
| Children (2–5 years) | 590 | 39.0 | 1.13 | 0.78–1.63 | 0.53 |
| **Enrolment location** |  |  |  |  |  |
| CHW | 40 | 2.7 | Ref. |  |  |
| PHC | 1471 | 97.4 | 0.57 | 0.17–1.91 | 0.36 |
| **Health Zone** |  |  |  |  |  |
| Ipamu | 716 | 47.4 | Ref. |  |  |
| Kenge | 500 | 33.1 | 6.30 | 3.30–12.05 | <0.001 |
| Kingandu | 295 | 19.5 | 0.83 | 0.48–1.44 | 0.51 |
| **Danger signs** |  |  |  |  |  |
| No/Others | 271 | 17.9 | Ref. |  |  |
| Yes (general danger signs) | 1049 | 69.4 | 1.12 | 0.70–1.78 | 0.64 |
| Weakness (asthenia) | 68 | 4.5 | 1.16 | 0.45–2.98 | 0.76 |
| Unable to sit | 123 | 8.1 | 1.39 | 0.61–3.13 | 0.43 |
| **RAS administration** |  |  |  |  |  |
| Yes | 1291 | 85.4 | Ref. |  |  |
| No | 220 | 14.6 | 0.21 | 0.13–0.33 | <0.001 |
| **Referral delay** |  |  |  |  |  |
| >1 day / Not documented | 445 | 29.4 | Ref. |  |  |
| 0 – 1 day | 1066 | 70.6 | 0.95 | 0.65–1.41 | 0.81 |
| **Malaria test result (RHF)** |  |  |  |  |  |
| Positive | 1227 | 81.2 | Ref. |  |  |
| Negative / Not done | 284 | 18.8 | 0.07 | 0.04–0.11 | <0.001 |
| **Anaemia at arrival at RHF** |  |  |  |  |  |
| Severe anaemia ($\leq 5$ g/dl) | 785 | 52.0 | Ref. | | |
| No/mild anaemia/not done | 726 | 48.1 | 0.44 | 0.27–0.72 | 0.001 |
| **Other comorbidities** |  |  |  |  |  |
| No | 802 | 53.1 | Ref. |  |  |
| Yes | 709 | 46.9 | 2.36 | 1.62–3.44 | <0.001 |
| **Blood transfusion** |  |  |  |  |  |
| Yes | 775 | 51.29 | Ref. |  |  |
| No | 736 | 48.71 | 0.53 | 0.32–0.87 | 0.01 |

### Determinants of health status on day 28 (well versus still sick, among the 2120 survivors)

Table 6 displays the odds to be cured among the 2120 children still alive on Day 28, at the time of the home visit by CARAMAL staff. Of 2120 patients that were alive on day 28, 1846 (87.1%) were healthy and 274 (12.9%) were sick.

**Table 6.** Estimated associations between selected factors and the health status of feverish children 28 days after initial contact with the health system. N = 2120 alive on Day 28. OR = Odds Ratio. CHW = Community Health Worker. PHC = Primary Health Care. RHF = Referral Health Facilities. RAS = Rectal Artesunate. ACT = Artemisinin-based Combination Therapy. 95% CI = 95% Confidence Interval. Hb = Haemoglobin. NA = Not applicable (because not at RHF).

| Determinants | n | % | Adjusted OR | 95% CI | p-value |
| --- | --- | --- | --- | --- | --- |
| **Age** |  |  |  |  |  |
| Children (0–2 years) | 1198 | 56.5 | Ref. |  |  |
| Children (2–5 years) | 922 | 43.5 | 0.83 | 0.63–1.10 | 0.20 |
| **Health Zone** |  |  |  |  |  |
| Ipamu | 842 | 39.7 | Ref. |  |  |
| Kenge | 734 | 34.6 | 1.48 | 1.05–2.07 | 0.02 |
| Kingandu | 544 | 25.7 | 0.62 | 0.40–0.97 | 0.04 |
| **Danger signs** |  |  |  |  |  |
| Yes, according to iCCM | 1477 | 69.7 | Ref. |  |  |
| No/Others | 392 | 18.5 | 0.93 | 0.65–1.33 | 0.68 |
| Weakness (asthenia) | 103 | 4.9 | 1.07 | 0.57–2.01 | 0.83 |
| Unable to sit | 148 | 7.0 | 1.05 | 0.59–1.85 | 0.87 |
| **RAS administration** |  |  |  |  |  |
| No | 306 | 14.4 | Ref. |  |  |
| Yes | 1814 | 85.6 | 0.64 | 0.45–0.92 | 0.02 |
| **Injectable antimalarial** |  |  |  |  |  |
| No / NA | 928 | 43.8 | Ref. |  |  |
| Yes | 1192 | 56.2 | 1.03 | 0.67–1.59 | 0.89 |
| **Oral antimalarial given at RHF** |  |  |  |  |  |
| No | 996 | 47.0 | Ref. |  |  |
| Yes | 1124 | 53.0 | 1.08 | 0.68–1.72 | 0.74 |
| **Oral treatment given at discharge or prescribed** |  |  |  |  |  |
| No | 1432 | 67.6 | Ref. |  |  |
| Yes | 688 | 32.5 | 1.12 | 0.76–1.64 | 0.58 |
| **Malaria test result on day 28** |  |  |  |  |  |
| Negative / not done | 1279 | 60.3 | Ref. |  |  |
| Positive | 841 | 39.7 | 4.67 | 3.47–6.30 | <0.001 |
| **Severe anaemia (day 28)** |  |  |  |  |  |
| No anaemia/not done | 790 | 37.3 | Ref. |  |  |
| Anaemia (Hb<11g/dL) | 1330 | 62.7 | 2.01 | 1.46–2.77 | <0.001 |

It appears that there was no evidence of a significant decrease in odds of still being ill on day 28 for children aged 2 to 5 years compared to those of age 0 to 2 years (p=0.20). However, study health zones showed heterogeneity of association with the health status of children: While the odds of being sick were higher in Kenge HZ (aOR = 1.48, 95% CI 1.05–2.07) compared to Ipamu (Ref), the reverse was observed at Kingandu HZ (aOR = 0.62, 95% CI 0.40–0.97). Surprisingly, the presence of iCCM general danger signs, as well as of other danger signs was not associated with the health status on day 28, when compared to iCCM standard danger signs. In addition, patients who have received RAS were less likely to be sick on day 28 (aOR = 0.64, 95% CI 0.45–0.92) compared to those who did not receive RAS. Injectable antimalarial treatment, oral antimalarial given at the RHF and a prescription for an oral antimalarial were not significantly associated with health status of patients on day 28. Patients that were tested positive for malaria on day 28 were significantly more likely to be sick at that time point (aOR = 4.67, 95% CI 3.47–6.30), and this was also the case for those with anaemia (Hb<11g/dL) (aOR = 2.01, 95% CI 1.46–2.77).

#### Determinants of death within 28 days after enrolment

In total, 161 enrolled children were deceased by the time of the Day 28 visit (Case Fatality Rate: 161 / 2281= 7.1 %). The great majority (n=137 or 85.1%) of these children had displayed standard iCCM danger signs at enrollment. 24 children had other danger signs (see Table 2 for list). The results shown in Table 7 focus only on the 161 children that passed away after having had danger signs at enrollment. Because “weakness” (n=103) was shown not to be a predictor of death, these 103 children were excluded, resulting in 2178 children analysed.

**Table 7.**
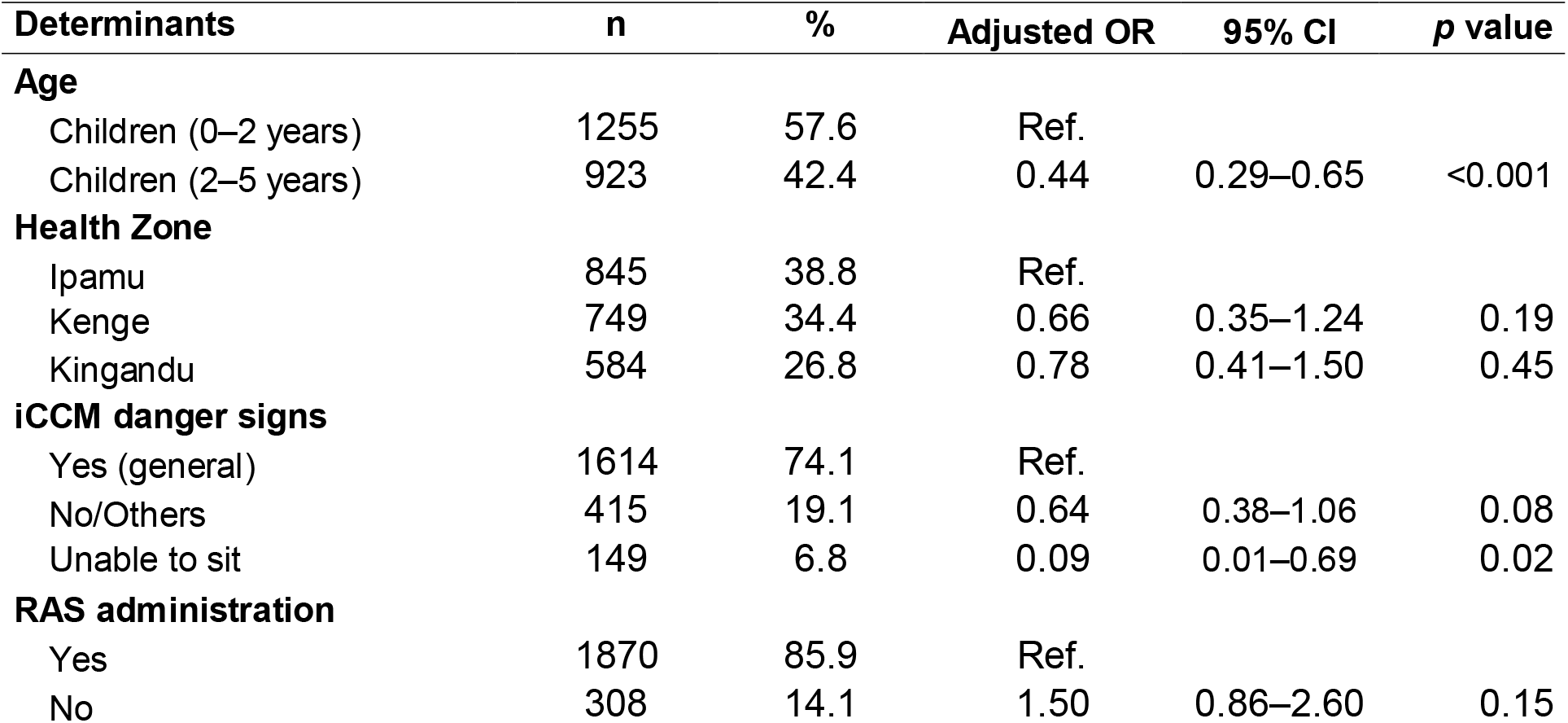
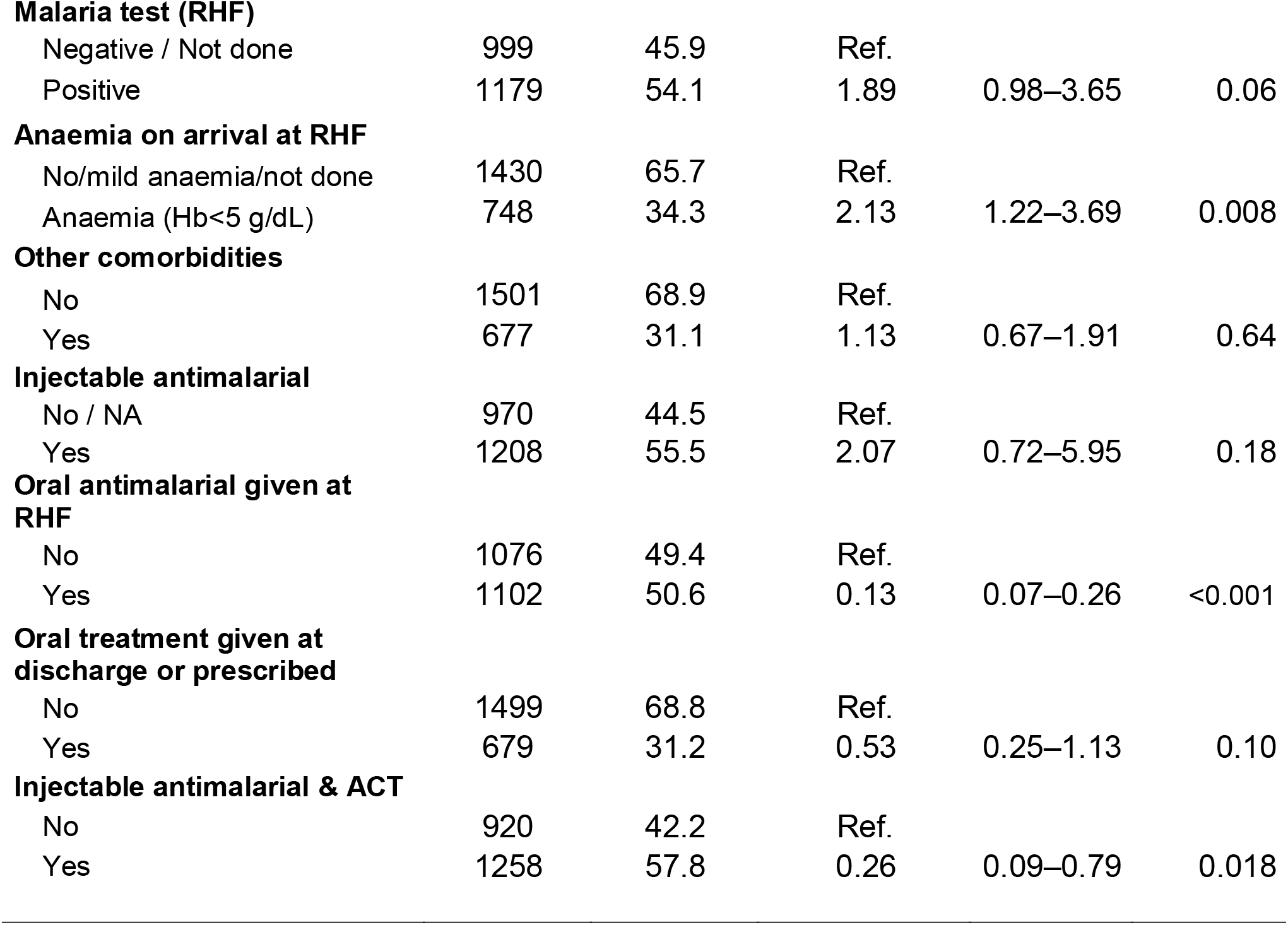
Determinants of death within 28 days following enrollment. N = 2178. OR = Odds Ratio. CHW = Community Health Worker. iCCM = integrated Community Case Management. PHC = Primary Health Care Center. RHF = Referral Health Facility. RAS = Rectal Artesunate. 95% CI = 95% Confidence Intervals. ACT = Artemisinin-based Combination Therapy. Hb = Haemoglobin. NA = Not applicable.

| Determinants | n | % | Adjusted OR | 95% CI | p value |
| --- | --- | --- | --- | --- | --- |
| **Age** |  |  |  |  |  |
| Children (0–2 years) | 1255 | 57.6 | Ref. |  |  |
| Children (2–5 years) | 923 | 42.4 | 0.44 | 0.29–0.65 | <0.001 |
| **Health Zone** |  |  |  |  |  |
| Ipamu | 845 | 38.8 | Ref. |  |  |
| Kenge | 749 | 34.4 | 0.66 | 0.35–1.24 | 0.19 |
| Kingandu | 584 | 26.8 | 0.78 | 0.41–1.50 | 0.45 |
| **iCCM danger signs** |  |  |  |  |  |
| Yes (general) | 1614 | 74.1 | Ref. |  |  |
| No/Others | 415 | 19.1 | 0.64 | 0.38–1.06 | 0.08 |
| Unable to sit | 149 | 6.8 | 0.09 | 0.01–0.69 | 0.02 |
| **RAS administration** |  |  |  |  |  |
| Yes | 1870 | 85.9 | Ref. |  |  |
| No | 308 | 14.1 | 1.50 | 0.86–2.60 | 0.15 |
| **Malaria test (RHF)** |  |  |  |  |  |
| Negative / Not done | 999 | 45.9 | Ref. |  |  |
| Positive | 1179 | 54.1 | 1.89 | 0.98–3.65 | 0.06 |
| **Anaemia on arrival at RHF** |  |  |  |  |  |
| No/mild anaemia/not done | 1430 | 65.7 | Ref. |  |  |
| Anaemia (Hb<5 g/dL) | 748 | 34.3 | 2.13 | 1.22–3.69 | 0.008 |
| **Other comorbidities** |  |  |  |  |  |
| No | 1501 | 68.9 | Ref. |  |  |
| Yes | 677 | 31.1 | 1.13 | 0.67–1.91 | 0.64 |
| **Injectable antimalarial** |  |  |  |  |  |
| No / NA | 970 | 44.5 | Ref. |  |  |
| Yes | 1208 | 55.5 | 2.07 | 0.72–5.95 | 0.18 |
| **Oral antimalarial given at RHF** |  |  |  |  |  |
| No | 1076 | 49.4 | Ref. |  |  |
| Yes | 1102 | 50.6 | 0.13 | 0.07–0.26 | <0.001 |
| **Oral treatment given at discharge or prescribed** |  |  |  |  |  |
| No | 1499 | 68.8 | Ref. |  |  |
| Yes | 679 | 31.2 | 0.53 | 0.25–1.13 | 0.10 |
| **Injectable antimalarial & ACT** |  |  |  |  |  |
| No | 920 | 42.2 | Ref. |  |  |
| Yes | 1258 | 57.8 | 0.26 | 0.09–0.79 | 0.018 |

Table 7 shows the associations between selected factors and the risk of dying within 28 days, after first contact with a CHW or PHC. Determinants of death within 28 days following enrollment was arguably the most important outcome in the CARAMAL study.

Compared to children between 0 to 2 years old as reference age group, children aged 2 to 5 years old were less likely to die (aOR = 0.44, 95% CI 0.29–0.65). Enrollment in the three study health zones was not found to be associated with death. The odds of dying were slightly lower but not significantly different between children not presenting iCCM general danger signs and those who had presented iCCM general danger signs (aOR = 0.64, 95% CI 0.38–1.06). However, the odds of dying were significantly lower among children “unable to sit” compared to those who had presented iCCM general danger signs (aOR = 0.09, 95% CI 0.01–0.69). The odds of dying was 1.50 times higher in patients that did not received RAS but the difference was unfortunately not significant, with a rather wide confidence interval (95% CI 0.86–2.60). Likewise, a positive malaria test at the RHF was a predictor of death, also the lower bound of the confidence interval was close to 1 (aOR = 1.88, 95% CI 0.97–3.62). In contrast, patients that were found with anaemia upon arrival at the RHF (aOR = 2.13, 95% CI 1.22–3.69) were more likely to die. The presence of comorbidities other than anaemia was associated with a non-significant increase in odds of dying (aOR = 1.13, 95% CI 0.67–1.91).

Injectable treatment taken alone did not show any evidence of effect on reducing the odds of death (aOR = 2.07, 95% CI 0.72–5.95). By contrast, patients that were treated with an oral antimalarial including ACT or oral quinine during admission (aOR = 0.13, 95% CI 0.07–0.26) were significantly less likely to die. Provision at discharge or prescription of an oral antimalarial treatment to be taken at home did show evidence of decreased odds of dying, but the result was not statistically significant (aOR = 0.53, 95% CI 0.25–1.13). Finally, patients that were treated with both an injectable antimalarial and an ACT (aOR = 0.26, 95% CI 0.09–0.79) were significantly less likely to die compared to those that were not. This obviously points towards the importance of proper case management for severe malaria cases.

## Discussion

In the CARAMAL study, the recognition of danger signs and symptoms of severe febrile illness by community-based providers (CHW and PHC) was the starting step for enrolling a child in the study. Firstly, this allowed to assess and classify sick children according to the iCCM or IMCI algorithms (11, 12). Secondly, it allowed to identify the proper course of action for the child, including immediate treatments and particularly the administration of RAS coupled with a recommendation for referral. While the evaluation of the overall effectiveness of RAS is the topic of another publication (28), here we tried here to investigate the value of a number of danger signs and other factors in view of key case management outcomes, and child mortality.

In DRC, some danger signs were used that are not part of the traditional iCCM general danger signs. Findings from this study suggest that the most frequently reported alternative danger sign was “unable to sit or stand up” (26.1%), which is similar to “unusually sleepy or unconscious at peripheral level”. Other alternative danger signs include “difficulty in breathing” (9.2%) and “white palms or sole” (6.8%), potential signs of severe anemia. Of note, most iCCM general danger signs appeared to increase during the post-RAS phase compared to the baseline phase. This could be the results of community sensitization and training of health workers prior to RAS rollout, which were held throughout the study settings by health authorities with support of UNICEF, just. Unfortunately, we have no independent measure to confirm this.

Little is known in the scientific literature on the frequency and importance of danger signs and how they predict RAS provision, referral, subsequent case management at a RHF, and ultimately the child’s health outcomes. These are some crucial points addressed by the CARAMAL project. In an earlier multi-country cluster randomized controlled trial conducted in Ghana, Guinea-Bissau, Tanzania and Uganda using pre-referral RAS at community level, (33), the odds ratio of being treated with RAS when a child presented danger signs was 1.84 (95% CI 1.20-2.83); p = 0.005). These findings are consistent with our results showing that those who presented iCCM general danger signs were significantly more likely to receive RAS (aOR = 2.77, 95% CI 2.04–3.77). Likewise, the trend was the same for the two additional signs triggering RAS use in DRC, although the association was not significant in children suffering from asthenia (weakness). Findings from Liberia have shown that the proportions of correct diagnosis and lifesaving treatment varied, especially for uncomplicated disease, but that the variability was lower for more severe cases, even though the accurate recognition of danger signs was sub-optimal (34).

As might be expected, iCCM general danger signs were a key determinant of RAS use in 66% of the enrolled children. In addition, two other signs triggering RAS use in DRC contributed to the total rate of children treated with RAS. RAS use was also affected by stock-outs of RAS and limited supplies at primary level (only 2 RAS doses per health worker at each replenishment). Heterogeneities in RAS use was observed between the three study Health Zones, due to differences in the availability of RAS (leading to more or less stock-outs), leadership of both HZ and PHCs, CHW and PHC coverage, and finally also health provider’s skills. Throughout the study implementation period, Kingandu HZ had consistently less stock outs of essential commodities including RAS, injectable drugs and ACT. It experienced few changes in leadership compared to the other two HZ, and this might be a reason for such good operational results. The latter is consistent with findings from a study conducted in Malawi that found that using RAS has the potential to increase utilization of child health services, but that this depended on the provider’s skills and their availability in remote areas(35). Very few caretakers refused the use of RAS (n=28), hence provider acceptabiity was not a problem in our setting.

One of the main purposes of RAS is to allow a safer referral, since lower level health facilities and CHW are not supposed to use injectable antimalarials. Hence, CARAMAL investigated referral determinants in detail. Overall referral completion in our study was 66.4%, associated with young age (children aged 0-2 years), PHC enrolments and RAS administration. The fact that infants were seen at a higher risk of complications would explain at least in part why younger children had higher referral rates. A reverse result was found in Uganda (36). RAS administration was significantly associated with increased odds of completing referral, which is similar with result from another study in Uganda(37).This may be explained in our study by the sensitization of caretakers and health worker during trainings prior to RAS rollout. However, our findings were different from another study in Uganda, which found that nearly all children treated with pre-referral RAS failed to comply with referral (38). Transport did not show an association with referral completion; eight in ten caretakers went by foot to a RHF, which was consistent with results from a study conducted in Afghanistan, where the majority of caretaker successfully completed referral by foot (39). Surprisingly, no evidence was found for an association between referral completion and presence of iCCM general danger signs. This does not match with evidence from another study (36). Additional factors based on our experience and reported in the literature are logistics, finances of the patients, communication skills, perceived quality of care, lack of time and need to care for other children and an improvement in the child’s condition (36, 38, 40, 41).

Injectable antimalarial treatment, especially of artesunate, is recommended in the national treatment guidelines to treat severe malaria at RHF. Treatment should continue for at least the first 24 hours, and continue until the patient becomes able to take oral medication (2, 14). In our study, the absence of association between iCCM general danger signs and injectable antimalarial treatment means a potential departure from the guidelines (2, 14) and raises a concern that severely ill children might not be treated in RHF appropriately to their condition.

However, given that RAS can promptly improve the child condition, and since 2 out of 3 children that received RAS also completed referral, it is well possible that some of these children improved on their way to a RHF, and hence were not eligible anymore for injectable antimalarials. Unfortunately, we could not follow the clinical conditions of the enrolled children in sufficient detail to investigate this.

Our findings suggest that injectable treatment alone did not seem to improve the health condition of the child. This is an important finding, which was also documented in the two other CARAMAL countries (Nigeria and Uganda). However, a reverse result was found in Liberia (42) where injectable artesunate was associated with a 78% reduction in hospital mortality (aOR = 0.22, 95% CI 0.07–0.67) and neurologic deficit in children of all ages.

By contrast, oral antimalarial treatment including an ACT or oral quinine while admitted in a RHF was significantly associated with a large decrease in the odds of dying – by 87%. The same strong effect was seen for the combination of an oral and a parenteral malaria treatment

– a 74% reduction in the risk of dying.

Severe anaemia upon arrival at RHF was significantly associated with more than doubling the odds of dying. Severe anemia is frequent in the DRC, and the lack of access to blood transfusion in remote areas (because of high cost and absence of blood banks) is a major issue. This result is similar to other found elsewhere in sub-Saharan Africa (43).

Finally, the odds of dying were 1.50 times higher in patients that did not received RAS, although the difference was on the margins of statistical significance. At least, this result provided some indication that RAS provision might be beneficial in this setting, and it was in the same order of effect as the results from the only randomized clinical trial of RAS done in Tanzania and Ghana (4). Here there results were obtained in the frame of a real-world setting, and are hence more likely to be generalizable. Our results on RAS mortality impact as not as good as the impact seen in another small study in Zambia (44). However, in that setting many other health system factors were optimized besides the provision of RAS.

### Limitations of the study

As with any observational study design, this study had some design limitations. The analysis presented here focuses on an individual patient analysis, for which many indicators were collected. To some extent, relevant confounders could therefore be controlled in the multivariate analysis, but it was impossible to avoid residual confounding. A major limitation was that despite our best efforts, it was difficult to track the clinical condition of the children for 28 full days, most of which being spent outside health facilities. Our field staff did their best to re-construct the treatment seeking pathway during the Day 28 interview at the home of the patient, focusing on issues such as location of care, treatment received, referrals. We are aware that a Day 28 interview bears a risk of rectal bias, and despite our best efforts through training and supervision, this certainly occurred. We then consolidated these results with the observations from enrollment and data from the RHF, if the children were brought there. But obviously, this still left some large gaps because the use of multiple providers, public and private, was the norm rather than the exception (Signorell *et al*., manuscript in preparation). In terms of methodological limitations, haemoglobin levels were measured with different methods across RHFs, resulting in minor difference of values depending on accuracy of each method used. Sahli’s hemoglobinometers were predominant at the beginning of the study, and were replaced progressively in all RHF by HemoCue photometer (HemoCueHb201+, Ängelholm, Sweden). A problem detected during the early course of the study was that initially we used mRDT that only assessed HRP2. Unfortunately, this molecule persists for weeks after an infection, hence it could not assess reliably parasitological cure at Day 28. Early on, the study switched to the combined HRP2/pLDH test, which was less prone to false positives after treatment. Lastly, we did not collect enough sociodemographic characteristics data related to parent’/caregiver’ because of the circumstances of the study. Data on socio-economic status would certainly have been important to include in our analysis.

## Conclusion

Our study aimed to describe key elements of case management for suspected severe cases of malaria, as well as the distribution of signs and symptoms among children <5 years. We investigated the differences in case management of children <5 years with different danger signs and varying treatment pathways, and related these to sickness and mortality outcomes. RAS seemed to offer some protections against dying, but it was not statistically significant.

Current definitions of danger signs in the DRC are rather un-standardized and potentially confusing, and the fact that DRC indicators are not standardized with global clinical algorithms for iCCM general is a problem. The lack of standardization of the signs and symptoms of severity made it also more difficult to estimate correctly the burden of severe febrile illness or severe malaria at community level.

Finally, proper case management at the RHF was greatly protective, and this constitutes an important study finding. They point once more to the importance of strengthening the health system in order to achieve both malaria control and child survival objectives.

## Data Availability

The datasets used and/or analysed during the current study are available from the corresponding author on reasonable request.

## List of abbreviations

95% CI: 95% Confidence Intervals
ACT: Artemisinin-based Combination Therapy
aOR: adjusted Odds Ratio
CARAMAL: Community Access to Rectal Artesunate for Malaria
CFR: Case Fatality Ratio
CHCS: Community Health Care Site
CHW: Community Health Worker
DHS-DRC II: DRC second Demographic and Health Survey
DRC: Democratic Republic of the Congo
Hb: Haemoglobin
*HRP2*: *Plasmodium falciparum* antigen histidine rich protein 2
HZ: Health Zone
iCCM: integrated Community Case Management
IQR: Interquartile Range
ITT: Intention-to-Treat
mRDT: malaria Rapid Diagnostic Test
NA: Not Applicable
ODK: Open Data Kit platform
OR: Odds Ratio
PHC: Primary Health Facility
pLDH: Plasmodium lactate dehydrogenase
PSS: Patient Surveillance System
RAS: Rectal Artesunate
RHF: Referral Health Facility
SD: Standard deviation
Swiss TPH: Swiss Tropical and Public Health Institute
UNICEF: United Nations Children’s Fund
WHO: World Health Organization

## Declarations

### Competing interests

All authors declared not having any financial relationships with any organizations that might have an interest in the submitted work in the previous three years, no any other relationships or activities that could appear to have influenced the submitted work.

### Funding

The CARAMAL Project was funded by Unitaid (grant reference XM-DAC-30010-CHAIRAS). The funder had no role in study design, data collection and analysis, decision to publish, or preparation of the manuscript.

## Acknowledgements

The authors would like to express their warm thanks to the children and parents/care givers who agreed to participate in the CARAMAL study, the health workers, local, provincial and national health authorities who provided their support especially the National Malaria Control Programme. A special thanks to the following individuals who contributed important aspects to the present work: Marek Kwiatkowski, Nadja Cereghetti (Swiss Tropical and Public Health Institute). Lydia Kabamba, Francine Kimanuka, Tony Byamungu (UNICEF DRC). Jenny Bokanga, Juliet Nakiganda, Carine Olinga (CHAI DRC). Jean-Claude Tembele (PNLP, DRC). Albert Kadjunga (PNECHOL-MD DRC). Ruffin Tuzolana, Louis Longa, Albert Caleb Koyelongo, Armand Mutwadi, Eddy Nzungu (Kinshasa School of Public Health, DRC). Yam’s Kabeya, Theodor Muamba (University of Kinshasa, DRC). Theodoor Visser, Harriet Napier (CHAI New York).

## Notes

### Competing Interest Statement

The authors have declared no competing interest.

### Author Declarations

The CARAMAL study protocol was approved by the Research Ethics Review Committee of the World Health Organization (WHO ERC, No. ERC.0003008), the Ethics Committee of the University of Kinshasa School of Public Health (No. 012/2018), the Health Research Ethics Committee of the Adamawa State Ministry of Health (S/MoH/1131/I), the National Health Research Ethics Committee of Nigeria (NHREC/01/01/2007-05/05/2018), the Higher Degrees, Research and Ethics Committee of the Makerere University School of Public Health (No. 548), the Uganda National Council for Science and Technology (UNCST, No. SS 4534), and the Scientific and Ethical Review Committee of CHAI (No. 112, 21 Nov 2017). The study is registered on ClinicalTrials.gov (NCT03568344). Consent was obtained in a two-step process given that the enrolled children were medical emergencies: a first provisional oral consent was obtained at the point of recruitment. The final written informed consent was then obtained during the first contact of the patient/caretaker with the study team - at the referral facility or during the day-28 home visit.

